# Apathy, dementia risk and mortality: long-term associations in a population-based prospective cohort of older adults

**DOI:** 10.1101/2025.03.03.25323298

**Authors:** Fleur Harrison, Moyra E. Mortby, Andrew R. Lloyd, Adam J. Guastella, Julian N. Trollor, Perminder S. Sachdev, Henry Brodaty

**Author notes:** **Corresponding author details:** Name: Fleur Harrison Postal address: CHeBA, School of Psychiatry, Level 1, AGSM Building (G27), UNSW SYDNEY NSW 2052, AUSTRALIA.

## Abstract

**Background:** Understanding whether apathy in older adults is related to incident dementia and mortality could help identify at-risk individuals, and inform public health efforts. This study aimed to investigate associations between apathy and these outcomes over long-term follow-up, and their independence from the overlapping symptoms of depression and fatigue.

**Methods:** In an Australian population-based cohort of 1,030 community-dwelling older adults aged 70-90, without dementia at baseline, apathy was assessed using the self-report Geriatric Depression Scale-3A subscale. Incident dementia was established via consensus diagnosis over 12 year follow-up, and mortality by record linkage over 18 years. We calculated hazard ratios (HRs) using Cox proportional hazards analyses. We repeated analyses adjusting for depression, fatigue and covariates, accounting for competing risk of mortality, and excluding short-term cases.

**Findings:** Unadjusted primary analyses showed the presence of self-reported apathy was associated with higher risk of dementia (HR 1.45, 95% confidence interval (CI) 1.05–2.00) and mortality (HR 1.76, 95% CI 1.43–2.16). Participants with apathy developed dementia a year earlier, and died three years earlier. These findings remained significant when adjusting for depression. The association with dementia was no longer significant when adjusting for fatigue or covariates, nor when taking mortality into account or excluding those cases where dementia developed in the shorter term.

**Interpretation:** The presence of apathy may represent an important risk indicator for dementia and mortality in older adults without dementia, independent of depression. Its association with dementia may reflect reverse causality. Future studies are needed to better understand the causal relationships that may underpin this observed association in the short and long-term, and the utility of apathy for screening in public health settings.

**Funding:** Dementia Australia Research Foundation, Centre for Healthy Brain Ageing.

## Introduction

Neuropsychiatric symptoms are core features of neurocognitive disorders, and almost universally experienced during the disease course^1^. Symptoms can initially emerge in otherwise healthy older adults, where they are associated with faster cognitive and functional decline, more amyloid and tau pathology, and higher conversion to dementia^2–4^. Neuropsychiatric symptoms may therefore represent early manifestations of dementia. This may be particularly true when symptoms emerge de novo and persist over time, as reflected by the new construct of mild behavioural impairment (MBI)^5^. The most prevalent and persistent among these symptoms is apathy, defined as quantitative reduction in motivation or goal-directed behaviour^6,7^. Apathy strongly impairs functional abilities, more so than other symptoms^7,8^, and increases risk of frailty, falls, cardiovascular disease, and costs of care ^9–13^. It has been linked with cognitive decline, and appears to increase risk of incident dementia, among persons with mild cognitive impairment (MCI)^13–15^.

Apathy thus holds substantial promise as a prevalent, easily measureable marker of poorer outcomes in older adults, which could identify at-risk individuals early, and inform efforts at risk reduction^14^. However, there are considerable gaps in evidence supporting this, particularly in population-based samples of older adults^16^. This lack of evidence could be because apathy conceptually overlaps with depression, and can be misdiagnosed as such^17^. Unlike depression, apathy is not an established psychiatric diagnosis according to the DSM-V, and is less well known^18^. Although these two symptoms can co-occur, they are increasingly seen as distinct entities or syndromes, differing in clinical profile, underlying neurobiology and potential treatments^19^. It is important to understand their differential effects, and evaluate their independence. However, most epidemiological research has considered them alone, not in conjunction, precluding their comparison. Moreover, apathy is included in some commonly-used measures of depression. So too is fatigue, which presents similarly to apathy and can also be considered an independent syndrome^20–22^. As a result, the use of standard depression sum-scores inadvertently masks the effects of individual symptoms such as apathy and fatigue, and may inflate the effect of depression^23^.

Reflecting these issues, there may be fifty epidemiological studies on depression in cognitively normal older adults as a risk factor for dementia^24–26^, but only six on apathy were identified in 2021 by systematic review^27^. Their pooled findings indicated apathy conferred at least two-fold risk for dementia; however, this was not the case in sensitivity analyses which adjusted for depression, or assessed apathy with a self-report (as opposed to informant-report) tool. Similarly, only two population-based studies have linked apathy with mortality, suggesting up to two-fold risk^13,28^, whereas depression is firmly established as a risk factor^29^. More epidemiological research is required to better understand the long-term impacts of apathy.

Along with the limited evidence base, and issues due to overlap with depression and fatigue, other limitations of the epidemiological literature on apathy include a) short follow-up periods (all less than ten years, the period considered sufficient by the Lancet Standing Commission^24^ to reduce likelihood of reverse causality), b) lack of adjustment for the competing hazard of mortality, and c) relatively young cohorts (up to 80 years). Most studies used informant-reported measures of apathy, which have not been validated for use in pre-dementia populations, and may not be comparable to self-reported measures, potentially producing different findings^30,31^. For persons without dementia, self-reported measures may be preferable as they reflect lived experience and are more sensitive^32^. Future research addressing these limitations is warranted.

Our primary aim is to investigate whether self-reported symptoms of apathy in community-dwelling older adults increase the hazard of incident dementia and mortality over long-term follow-up. We address the key issue of overlapping symptoms by reporting findings for apathy, depression and fatigue alone, in order to compare their effects, and then after adjustment for the other two symptom, to evaluate their independence. We test the robustness of apathy findings in various sensitivity analyses. These include accounting for the competing risk of mortality, and excluding short-term cases to minimise reverse causality. The secondary aim is to determine the impact of using different apathy measures on study findings.

## Methods

### Study design and setting

#### Participants

The Sydney Memory and Ageing Study (MAS) was designed as a population-based epidemiological cohort of community-dwelling adults, aged between 70 and 90 and without dementia. All individuals in a defined area of Sydney, NSW, Australia, based on mandatory enrolment on the electoral roll, received a mail-out invitation to participate. 1,037 persons met inclusion/exclusion criteria, provided informed written consent, and undertook a comprehensive in-person baseline assessment, between 2005-2007. Full details on exclusion criteria are provided in Sachdev et al.^33^, but in short, excluded dementia and neurodegenerative conditions at baseline. Optional assessments included fasting blood or saliva collection, MRI scan and informant phone interview. Follow-up assessments took place every 2 years, for 12 years unless participants withdrew or died. Ethics approval was obtained from UNSW Sydney and South-Eastern and Illawarra Area Health Service committees.

### Measures

#### Apathy, depression and fatigue

Self-report apathy and depression measures were obtained from the 15-item Geriatric Depression Scale^34^, by creating two subscales in line with previous studies^10,11,13,28^. Items are scored as present or absent over the last week, and do not include somatic or sexual symptoms. The apathy subscale (GDS-3A) comprises three items (2: ‘Have you dropped many of your activities and interests?’; 9: ‘Do you prefer to stay home at night, rather than going out and doing things?’; 13: ‘Do you feel full of energy?’ reverse coded). Responses are summed for total score (range 0-3). Evidence supporting this subscale includes: substantial evidence of a separate apathy factor, including from meta-analysis of 26 factor analytic studies^35^; clinical consensus on its face validity^36^; and reports of acceptable psychometric properties, including diagnostic accuracy^36,37^. Item 9 was slightly modified in MAS, following Brink’s original version^38^; the three items loaded on an apathy factor, irrespective of this modification^39^. The remaining 12 items were used to assess depression. This measure, termed the GDS-12D (range 0-12) has not been psychometrically evaluated, but is used consistently alongside the GDS-3A. Items cover symptoms such as low mood and affect, hopelessness, worthlessness and not wanting to be alive. Participants missing >1 item on the GDS3A or >2 on the GDS-12D were excluded from analyses. For primary analyses, we defined the presence of clinically relevant symptoms by using “strict” cutoff scores (GDS-3A=3; GDS-12D≥4). The former was based on diagnostic accuracy analysis^37^; the latter by pro-rating the well-established GDS-15 cutoff score (≥5). Secondary analyses used firstly, “standard” cutoffs (GDS-3A≥2; GDS-12D≥2) as used in previous research although having low sensitivity^13,36^, and secondly, continuous subscale scores.

Fatigue was assessed with item 12 of the Assessment of Quality of Life-6D (AQoL-6D; ‘How much energy do you have to do the things you want to do?’)^40^. After reverse-scoring and recoding the Likert-type response options, a higher score represented greater fatigue (range 0-4). This single-item fatigue measure had concurrent validity against more comprehensive measures of fatigue ^41^. For both primary and secondary analyses, the presence of clinically relevant fatigue was defined by responses “usually tired and lacking energy” or “always tired and lacking energy” (score ≥3), based on similar measures. A continuous fatigue score was also used in secondary analyses.

Informant-report apathy was from the NeuroPsychiatric Inventory (NPI)^42^. During phone interviews, if a screening question indicated the presence of apathy over the last four weeks, ratings of frequency (1-*occasionally* to 4-*very frequently*) and severity (1-*mild* to 3-*severe*) were multiplied to calculate the domain score (range 0-12). Although widely used, the NPI apathy domain has evidence of inadequate psychometric properties^30^. A cut-off score validated in a dementia-free sample (≥1) was applied.

### Dementia and mortality

The dementia outcome variable was time to clinical diagnosis. Diagnoses using DSM-IV criteria were available for all six waves of follow-up data collection, i.e. for up to 12 years. At each wave, expert clinicians from a panel of psychogeriatricians, neuropsychiatrists, clinical psychologists and neuropsychologists discussed all available information, including MRI where available, to reach a consensus diagnosis. For participants who received a diagnosis, time to diagnosis was calculated as time from baseline assessment to the midway point between the assessment when dementia was first diagnosed and the previous assessment. Participants who had complete neuropsychological test data and did not meet criteria for a dementia diagnosis were classified as “not having dementia” at each wave. For participants who never received a diagnosis, time to diagnosis was censored at the 12-year follow-up time point or drop-out from the study.

For all-cause mortality, the outcome variable was time to death. Death information was ascertained from the Australian National Death Index (NDI), up to December 2018. Where NDI data were not available, we relied on information obtained from Ryerson Index of Obituaries, relatives or friends, and hospital/nursing home staff. The most recent determination of deaths from Ryerson Index occurred in March 2024, when all remaining survivors were cross-referenced, and deaths recorded where there was an unambiguous match. Time to death was calculated as time from baseline assessment to death, or from baseline to March 1^st^, 2024 for survivors.

### Covariates

Participants reported age at initial assessment, sex, education, non-English-speaking background (NESB, defined as learning conversational English aged ≥10), history of diagnosed stroke or cardiovascular disease, and use of antidepressant medication. Global cognition was assessed with MMSE^43^. Apolipoprotein E (*APOE*) L4 genotype was assessed by blood or saliva DNA analysis.

### Analyses

Analyses were conducted in STATA and SPSS, with statistical significance set at *p*<.05. Descriptive analyses were *t*-tests and χ^2^, to investigate differences between participants with and without apathy at baseline. Participants with missing data were left out of analyses. Primary analyses were proportional hazard Cox regressions. Predictor variables were the presence of apathy, depression or fatigue (using strict cutoffs) at baseline. Endpoints were time dementia diagnosis or mortality. Model 1 was unadjusted; model 2 adjusted for depression (in apathy and fatigue analyses) or apathy (in depression analyses). Model 3 additionally adjusted for age, gender, education, NESB, baseline MMSE score, *APOE* L4 allele carriage, history of stroke/cardiovascular disease, and antidepressant use. Model 3 additionally adjusted for fatigue (in apathy and depression analyses) or apathy (in fatigue analyses). We tested all covariates for potential interactions with apathy symptoms, corrected for the number of interactions by Bonferroni correction.

We conducted several sensitivity analyses: (1) to minimise possible reverse causality for dementia, we examined HRs when excluding cases of dementia which occurred within four years’ of baseline, and visually examined whether the bivariate survival curves for participants with and without apathy diverged consistently over time; (2) to take the competing risk of mortality into account (as it could have precluded dementia from occurring), we used Fine-Gray subdistribution hazard models to estimate subhazard ratios (SHR); (3) to evaluate whether apathy is associated with endpoints in older rather than younger participants, we examined HRs using split-half groups by age; (4) to explore whether apathy is associated with endpoints independently of fatigue, we performed analyses with one fatigue-related item (item 13) removed from the apathy measure, which we termed the GDS-2A.

Secondary analyses were identical to primary analyses, other than the following variations to the predictor variables: A) using standard cutoffs on the apathy and depression GDS subscales; B) using continuous scores of apathy, depression and fatigue; C) using an informant-report apathy measure (NPI).

## Results

### Cohort characteristics

As shown in Table 1, 1,030 community-dwelling individuals aged 70-90 were included from the Sydney Memory and Ageing Study cohort at baseline (see Supplementary Figure 1). Compared to those without apathy, participants with apathy (GDS-3A=3) were older, had higher depression and fatigue scores, and were more likely to be taking antidepressant medication or have a history of stroke or cardiovascular disease. Other characteristics did not differ between the two groups.

**TABLE 1.**
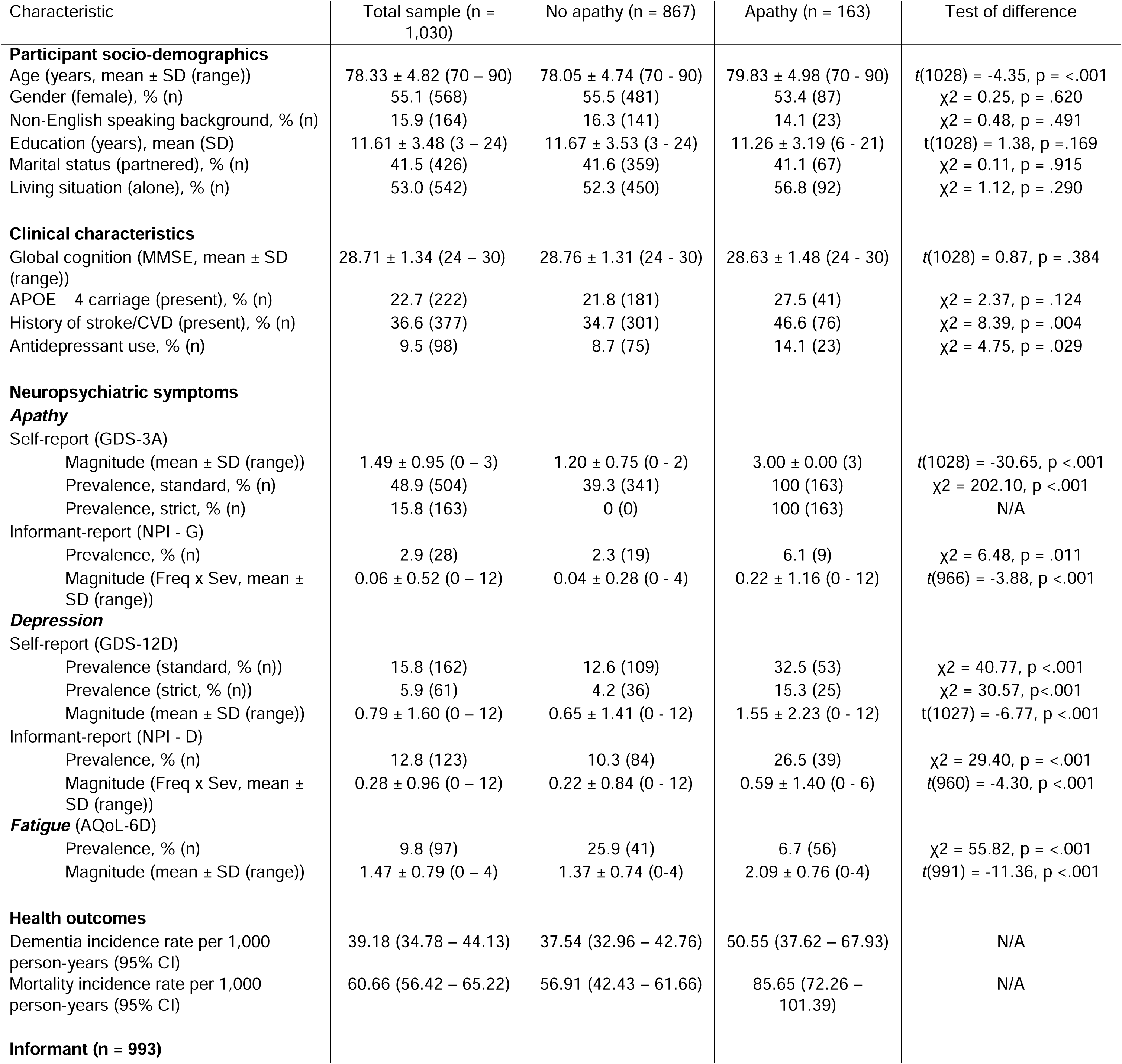
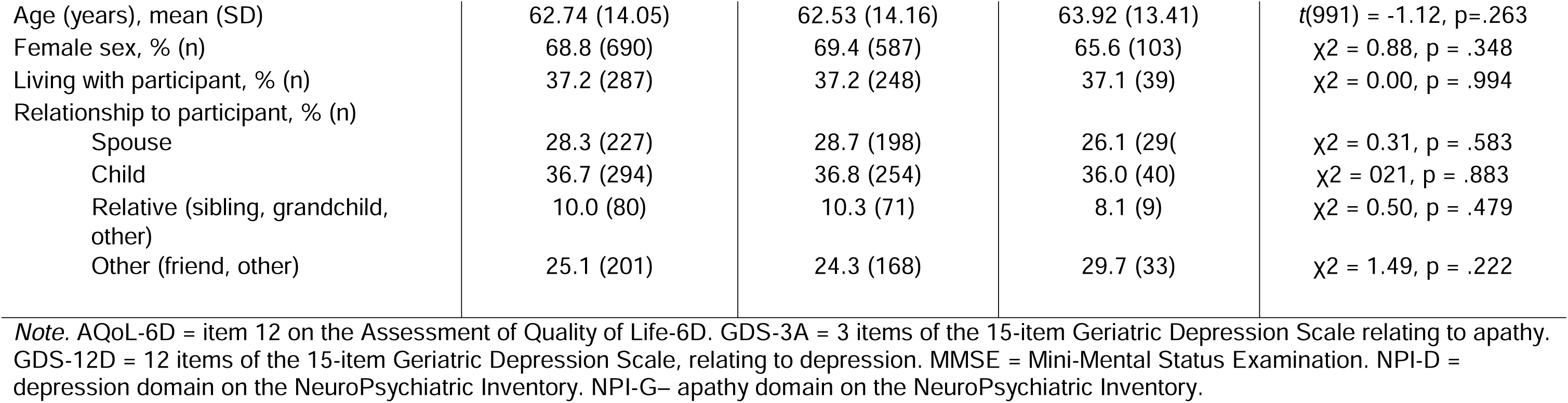
Baseline characteristics and health outcomes of the Sydney Memory and Ageing Study cohort, and according to presence of apathy (defined as GDS-3A =3)

| Characteristic | Total sample (n = 1,030) | No apathy (n = 867) | Apathy (n = 163) | Test of difference |
| --- | --- | --- | --- | --- |
| **Participant socio-demographics** |  |  |  |  |
| Age (years, mean ± SD (range)) | 78.33 ± 4.82 (70 – 90) | 78.05 ± 4.74 (70 - 90) | 79.83 ± 4.98 (70 - 90) | $t(1028) = -4.35, p = <.001$ |
| Gender (female), % (n) | 55.1 (568) | 55.5 (481) | 53.4 (87) | $\chi^2 = 0.25, p = .620$ |
| Non-English speaking background, % (n) | 15.9 (164) | 16.3 (141) | 14.1 (23) | $\chi^2 = 0.48, p = .491$ |
| Education (years), mean (SD) | 11.61 ± 3.48 (3 – 24) | 11.67 ± 3.53 (3 - 24) | 11.26 ± 3.19 (6 - 21) | $t(1028) = 1.38, p = .169$ |
| Marital status (partnered), % (n) | 41.5 (426) | 41.6 (359) | 41.1 (67) | $\chi^2 = 0.11, p = .915$ |
| Living situation (alone), % (n) | 53.0 (542) | 52.3 (450) | 56.8 (92) | $\chi^2 = 1.12, p = .290$ |
| **Clinical characteristics** |  |  |  |  |
| Global cognition (MMSE, mean ± SD (range)) | 28.71 ± 1.34 (24 – 30) | 28.76 ± 1.31 (24 - 30) | 28.63 ± 1.48 (24 - 30) | $t(1028) = 0.87, p = .384$ |
| APOE ε4 carriage (present), % (n) | 22.7 (222) | 21.8 (181) | 27.5 (41) | $\chi^2 = 2.37, p = .124$ |
| History of stroke/CVD (present), % (n) | 36.6 (377) | 34.7 (301) | 46.6 (76) | $\chi^2 = 8.39, p = .004$ |
| Antidepressant use, % (n) | 9.5 (98) | 8.7 (75) | 14.1 (23) | $\chi^2 = 4.75, p = .029$ |
| **Neuropsychiatric symptoms** |  |  |  |  |
| **Apathy** |  |  |  |  |
| Self-report (GDS-3A) |  |  |  |  |
| Magnitude (mean ± SD (range)) | 1.49 ± 0.95 (0 – 3) | 1.20 ± 0.75 (0 - 2) | 3.00 ± 0.00 (3) | $t(1028) = -30.65, p <.001$ |
| Prevalence, standard, % (n) | 48.9 (504) | 39.3 (341) | 100 (163) | $\chi^2 = 202.10, p <.001$ |
| Prevalence, strict, % (n) | 15.8 (163) | 0 (0) | 100 (163) | N/A |
| Informant-report (NPI - G) |  |  |  |  |
| Prevalence, % (n) | 2.9 (28) | 2.3 (19) | 6.1 (9) | $\chi^2 = 6.48, p = .011$ |
| Magnitude (Freq x Sev, mean ± SD (range)) | 0.06 ± 0.52 (0 – 12) | 0.04 ± 0.28 (0 - 4) | 0.22 ± 1.16 (0 - 12) | $t(966) = -3.88, p <.001$ |
| **Depression** |  |  |  |  |
| Self-report (GDS-12D) |  |  |  |  |
| Prevalence (standard, % (n)) | 15.8 (162) | 12.6 (109) | 32.5 (53) | $\chi^2 = 40.77, p <.001$ |
| Prevalence (strict, % (n)) | 5.9 (61) | 4.2 (36) | 15.3 (25) | $\chi^2 = 30.57, p <.001$ |
| Magnitude (mean ± SD (range)) | 0.79 ± 1.60 (0 – 12) | 0.65 ± 1.41 (0 - 12) | 1.55 ± 2.23 (0 - 12) | $t(1027) = -6.77, p <.001$ |
| Informant-report (NPI - D) |  |  |  |  |
| Prevalence, % (n) | 12.8 (123) | 10.3 (84) | 26.5 (39) | $\chi^2 = 29.40, p = <.001$ |
| Magnitude (Freq x Sev, mean ± SD (range)) | 0.28 ± 0.96 (0 – 12) | 0.22 ± 0.84 (0 - 12) | 0.59 ± 1.40 (0 - 6) | $t(960) = -4.30, p <.001$ |
| **Fatigue** (AQoL-6D) |  |  |  |  |
| Prevalence, % (n) | 9.8 (97) | 25.9 (41) | 6.7 (56) | $\chi^2 = 55.82, p = <.001$ |
| Magnitude (mean ± SD (range)) | 1.47 ± 0.79 (0 – 4) | 1.37 ± 0.74 (0-4) | 2.09 ± 0.76 (0-4) | $t(991) = -11.36, p <.001$ |
| **Health outcomes** |  |  |  |  |
| Dementia incidence rate per 1,000 person-years (95% CI) | 39.18 (34.78 – 44.13) | 37.54 (32.96 – 42.76) | 50.55 (37.62 – 67.93) | N/A |
| Mortality incidence rate per 1,000 person-years (95% CI) | 60.66 (56.42 – 65.22) | 56.91 (42.43 – 61.66) | 85.65 (72.26 – 101.39) | N/A |
| **Informant (n = 993)** |  |  |  |  |
| Age (years), mean (SD) | 62.74 (14.05) | 62.53 (14.16) | 63.92 (13.41) | $t(991) = -1.12, p = .263$ |
| Female sex, % (n) | 68.8 (690) | 69.4 (587) | 65.6 (103) | $\chi^2 = 0.88, p = .348$ |
| Living with participant, % (n) | 37.2 (287) | 37.2 (248) | 37.1 (39) | $\chi^2 = 0.00, p = .994$ |
| Relationship to participant, % (n) |  |  |  |  |
| Spouse | 28.3 (227) | 28.7 (198) | 26.1 (29) | $\chi^2 = 0.31, p = .583$ |
| Child | 36.7 (294) | 36.8 (254) | 36.0 (40) | $\chi^2 = 0.21, p = .883$ |
| Relative (sibling, grandchild, other) | 10.0 (80) | 10.3 (71) | 8.1 (9) | $\chi^2 = 0.50, p = .479$ |
| Other (friend, other) | 25.1 (201) | 24.3 (168) | 29.7 (33) | $\chi^2 = 1.49, p = .222$ |

*Note.* AQoL-6D = item 12 on the Assessment of Quality of Life-6D. GDS-3A = 3 items of the 15-item Geriatric Depression Scale relating to apathy. GDS-12D = 12 items of the 15-item Geriatric Depression Scale, relating to depression. MMSE = Mini-Mental Status Examination. NPI-D = depression domain on the NeuroPsychiatric Inventory. NPI-G = apathy domain on the NeuroPsychiatric Inventory.

### Long-term incident dementia and mortality

Follow-up data on dementia were available for 854/1030 (82.9%) participants, with mean follow-up time of 9.53 years (range 1.80-13.72). Compared to those followed up, participants without dementia follow-up data were older, had more chronic conditions, and greater apathy, depression and fatigue scores at baseline (Suppl Table 1). Among the 854 with follow-up data, 271 (31.7%) developed dementia, a mean of 10.92 years after baseline (95% CI 10.64–11.20). Those who did were older, more likely to carry an *APOE* L4 allele or be of non-English-speaking background, and had poorer cognition at baseline (Suppl Table 2).

Mortality data were available for 1,028/1,030 (99.8%) MAS participants. The majority died during the 18-year follow-up period (n=733, 71.3%). Mean survival time was 12.07 years (95% CI 11.74–12.41).

### Associations of apathy, depression and fatigue with incident dementia

Descriptively, in the apathy group, 36.1% of participants (44/122) developed dementia, a mean of 9.83 years after baseline (95% CI 9.06–10.61). This compared to 31.0% of participants without apathy (227/732), after mean 11.05 years (95% CI 10.75–11.34). Kaplan-Meier survival curves by apathy status indicated the proportional hazards assumption was adequately met (see Figure 2). This was also the case for depression and fatigue (Suppl Figures 2A and 3A).

**Figure 1.**
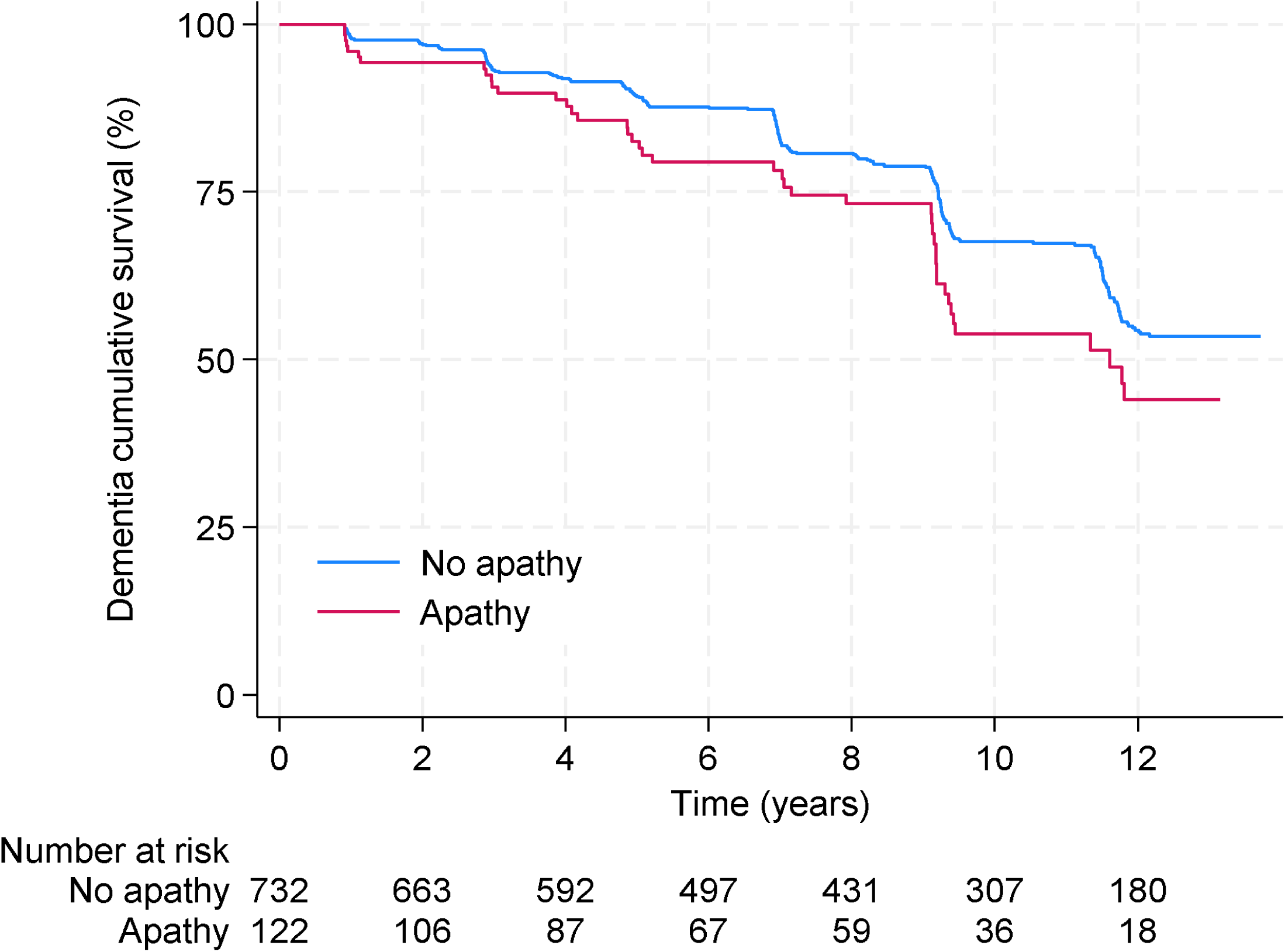
Proportion of participants without dementia over 12 years, for those with and without apathy (GDS-3A = 3) (χ*2* = 5.08, *p* =.024 by log rank test)

**Figure 2.**
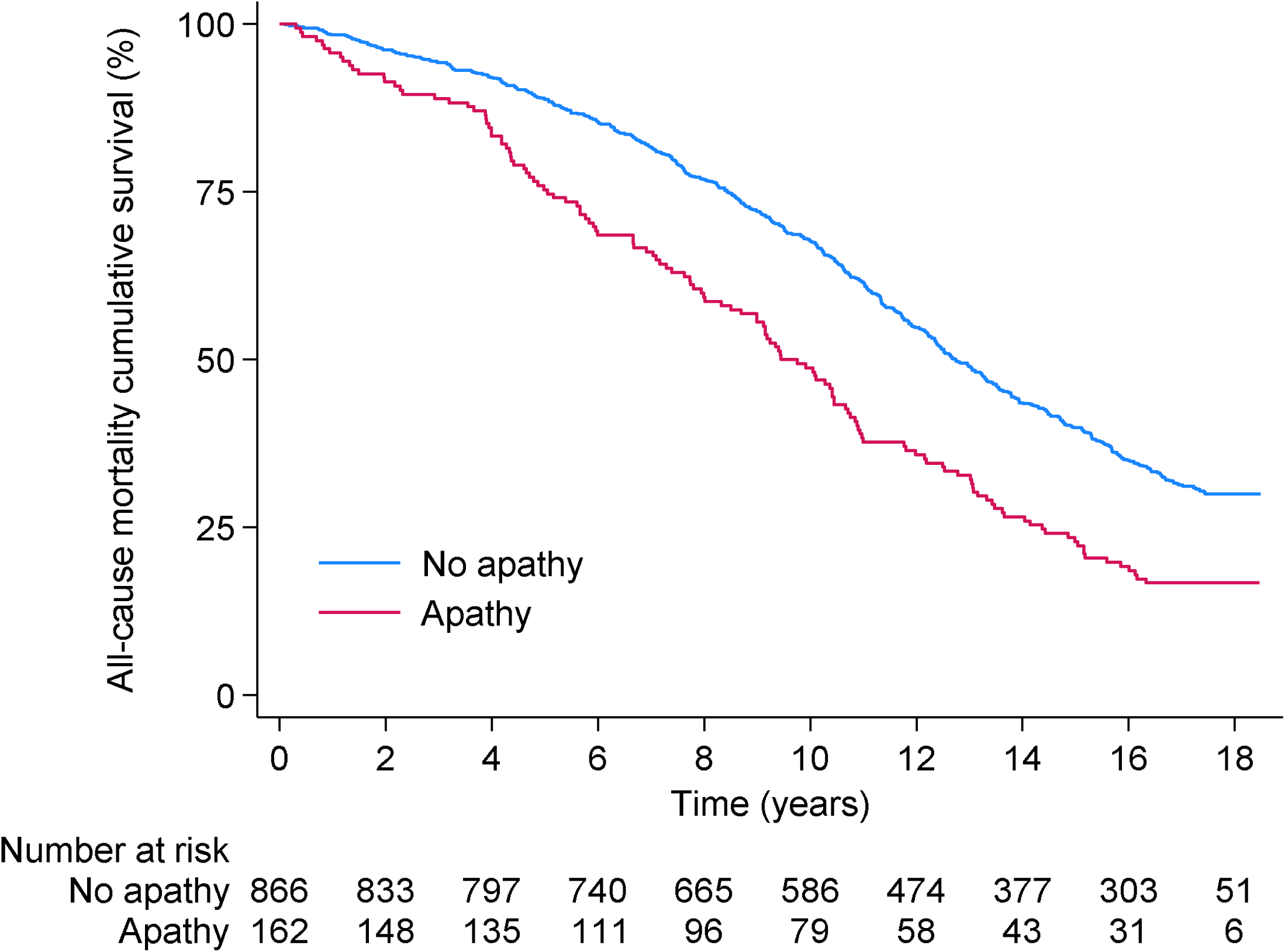
Proportion of survivors over 18 years, for participants with and without apathy (GDS-3A=3) (χ*2* =29.,31, *p* =<.001 by log rank test)

Results of the primary analyses are shown in Table 2. Apathy was significantly associated with higher risk of dementia, in the unadjusted model (HR 1.45, 95% CI 1.05 – 2.00, *p*=.025) and after adjusting for depression (Model 1). This association was no longer significant in the fully adjusted model (Model 2), and finally, when fatigue was also included (Model 3). There were no significant interactions for apathy with depression, fatigue or covariates in these analyses.

**TABLE 2.** Associations of baseline apathy, depression and fatigue with incident dementia and mortality.

| Dementia |  |  |  |  |  |  |  |  |  |  |  |  |  |
| --- | --- | --- | --- | --- | --- | --- | --- | --- | --- | --- | --- | --- | --- |
|  |  | Unadjusted |  |  | Model 1 |  |  | Model 2 |  |  | Model 3 |  |  |
|  | Event/total | HR | 95% CI | *p* value | HR | 95% CI | *p* value | HR | 95% CI | *p* value | HR | 95% CI | *p* value |
| Apathy | 271/854 | 1.45 | 1.05–2.00 | .025 | 1.44 | 1.04 – 2.00 | .030 | 1.17 | .83 -1.65 | .369 | 1.16 | .81 – 1.68 | .416 |
| Depression | 270/854 | 1.22 | .70–2.13 | .491 | 1.10 | .62 – 1.94 | .744 | 1.11 | .61 – 2.00 | .739 | 1.07 | .58 – 1.98 | .825 |
| Fatigue | 261/835 | 1.59 | 1.04–2.42 | .032 | 1.58 | 1.03 – 2.41 | .036 | 1.13 | .72 – 1.78 | .597 | 1.07 | .66 – 1.72 | .793 |
| Mortality |  |  |  |  |  |  |  |  |  |  |  |  |  |
|  |  | Unadjusted |  |  | Model 1 |  |  | Model 2 |  |  | Model 3 |  |  |
|  | Event/total | HR | 95% CI | *p* value | HR | 95% CI | *p* value | HR | 95% CI | *p* value | HR | 95% CI | *p* value |
| Apathy | 733/1,028 | 1.67 | 1.38 – 2.01 | <.001 | 1.58 | 1.30 – 1.91 | <.001 | 1.28 | 1.05 – 1.57 | .016 | 1.19 | .96 – 1.48 | .108 |
| Depression | 732/1,027 | 1.85 | 1.40 – 2.44 | <.001 | 1.61 | 1.21 – 2.14 | .001 | 1.64 | 1.22 – 2.21 | .001 | 1.63 | 1.20 – 2.23 | .002 |
| Fatigue | 710/995 | 1.87 | 1.49 – 2.35 | <.001 | 1.80 | 1.46 – 2.26 | <.001 | 1.41 | 1.10 – 1.80 | .006 | 1.35 | 1.05 – 1.73 | .020 |

Note. *HR* = Hazard ratio. 95% CI = 95% confidence interval. Model 1 adjusted for depression (in apathy and fatigue analyses) or apathy (in depression analysis). Model 2 additionally adjusted for sex, age at baseline, non-English speaking background, education, global cognition, *APOE* $\epsilon$ 4 allele carriage, antidepressant use, history of stroke or cardiovascular disease. Model 3 additionally adjusted for fatigue (in apathy and depression analyses) or apathy (in fatigue analysis).

Regarding other symptoms (also shown in Table 2), self-report depression did not predict incident dementia; self-report fatigue was associated with risk of dementia when unadjusted and adjusting for depression, but not when adjusting for covariates or fatigue.

We tested the robustness of the primary result for apathy, using four sensitivity analyses. First, apathy was not associated with incident dementia, when excluding dementia diagnosed in the first four years’ follow-up (unadjusted HR 1.43, 95% CI .98-2.09; Suppl Table 6). Second, apathy was not associated with incident dementia in competing hazard analyses accounting for mortality (unadjusted sub-hazard ratio (SHR) 1.24, 95% CI .89-1.72; Suppl Table 10). Third, results were evaluated in younger (aged <78) and older (aged ≥78) groups. In the younger group, apathy was not significantly associated with dementia, but fatigue was (Suppl Table 7). In the older group, the results were comparable with the main findings, in that apathy significantly predicted dementia when unadjusted, but not when fully adjusted (Suppl Table 8). Fourth, after removing the fatigue-related GDS item to form the GDS-2A, associations between apathy and dementia were less strong (Suppl Table 9).

Secondary analyses were run, to explore using different apathy measures to predict risk of dementia. When applying the standard cutoff to the self-report measure, or its continuous score, results differed from the primary result, in that apathy was no longer associated with risk of dementia (Suppl Tables 3 and 4). By contrast, when using the informant-report measure (NPI, Suppl Table 5), a significant association was apparent, but only when adjusting for covariates.

### Associations of apathy, depression and fatigue with mortality

In the apathy group, 83.3% died (135/162), compared to 69.1% of those without apathy (598/866). Mean survival time for participants with apathy was 9.92 years (95% CI 9.07-10.77), compared with 12.47 years for others (95% CI 12.12-12.83). Kaplan-Meier survival curves by apathy status, as shown in Figure 3, showed no violations of the proportional-hazard assumption (also the case for depression and fatigue; see Suppl Figures 2B and 3B).

In the primary analyses (Cox regressions using strict cutoffs on self-report measures), apathy significantly increased risk of mortality when unadjusted (HR 1.67, 95% CI 1.38–2.01, *p*<.001; Table 2), and after adjustment for depression and other covariates (Models 2 & 3). However, the association was attenuated when further adjusting for fatigue (Model 4). Depression and fatigue were independently associated with mortality in all models (also Table 2). There were no significant interactions between apathy and depression, fatigue or covariates.

Sensitivity analyses were undertaken, firstly for younger (<78 years) and older (≥78 years) participants (Suppl Tables 7 and 8). Apathy’s association with mortality was not robust to confounding in the younger group, but remained so in the older group. Secondly, using the modified apathy measure (GDS-2A) made no difference to its associations with mortality (Suppl Table 9).

Secondary analyses revealed that varying the apathy measure did not change its association with mortality. All three alternative measures (two variations on the self-report measure, and the informant-report measure) were significantly related to this outcome (Suppl Tables 3,4,5).

## Discussion

In this population-based cohort of older adults, key findings were that the presence of apathy according to a brief self-report measure conferred 45% higher risk for dementia over 12 years, and 67% higher risk of mortality over 18 years, when unadjusted for other factors. Participants with apathy at baseline developed dementia over a year earlier than those without, and died three years earlier. Building on previous studies, our results confirm the potential utility of this easily measureable symptom as a precursor to shorter healthspan and longevity^13,14,27^.

### Apathy is associated with greater long-term risk of dementia and mortality

However, the association with incident dementia was not clear-cut. Its strength and significance were attenuated when a) fully adjusted, b) when excluding short-term dementia cases (in the first 4 years of follow-up), c) when taking mortality into account, and d) in younger participants (<78 years). Nonetheless, the direction of the association was maintained in all analyses. These results diverge with findings from the two comparable studies which examined self-reported apathy. In these, up to 2-fold risk for dementia was reported, which a) was independent of covariates, b) did not change over time, and c) was not altered by the competing risk of mortality^28,44^. These studies, as well as others using informant-report measures^3,45–47^, differed from ours in a number of ways. They had shorter follow-up periods, younger participants, and all bar one lacked genotype data. Ours is the first to report follow-up data of at least ten years, and to cover a broad age range. We adjusted for *APOE* L4 status, accounted for the competing hazard of mortality, and explored the impact of varying the apathy measure.

Our results almost completely agree with the 2021 meta-analysis^27^, in which the pooled association was not significant in subgroups of studies a) with longer follow-up, b) of younger participants, or c) using a self-report measure. As such, the current research provides a valuable, nuanced perspective on the epidemiology of apathy. It implies that apathy is more related to the development of dementia in the short term, rather than the long term. This supports the hypothesis that apathy is a prodromal symptom of dementia, or that the relationship represents reverse causality. This study also highlights the potentially complex inter-relationships between apathy and dementia, which both may represent decades of accumulated risks, influenced by socio-demographic, clinical and biological characteristics. Finally, it reveals that mortality may occur before dementia develops, thus diminishing the strength of the association.

By contrast, we found very consistent results for the other primary outcome, mortality. Apathy was independently associated with risk of death, irrespective of the measure used, and this remained the case in sensitivity analyses. This is consistent with comparable research^13,28^, although extends the time frame of the relationship to almost two decades. Most previous research linking apathy with mortality is drawn from clinical samples, e.g. frailty, dementia or neurological conditions^48,49^, so the current finding provides important validation of its public health relevance in the general population.

### The associations of apathy with dementia risk and mortality are independent of depression, but not fatigue

The current research aimed to clearly differentiate symptoms of apathy from those of depression and fatigue. Controlling for depression had no impact on results in our survival analyses, consistent with the few studies which also did this^13,28,44^, and supporting the independence of the two constructs. However, it differs from the corresponding sensitivity analysis of the 2021 meta-analysis. Surprisingly, we found depressive symptoms as measured by the GDS-12D were largely unrelated to subsequent dementia. This is contrary to strong evidence that depression is an important risk factor for dementia, according to “meta-umbrella” and umbrella reviews^25,26^, with 3% of dementia incidence theoretically preventable by removing risk due to depression^24^. However, understanding of this complex relationship remains incomplete^50^. Importantly, depressive disorders are highly heterogenous, yet typically measured by a single sum score, meaning important differences between symptoms may be masked^23^. An alternative approach is to consider individual symptoms or clusters. As shown here and previously^28^, doing so reveals that dementia risk typically attributed to depression may, in fact, be driven by questionnaire items on apathy, fatigue, and/or memory complaints, rather than mood symptoms. This warrants further investigation. Reverse causation may also play a role. On the other hand, depressive symptoms were independently associated with mortality, in this and two prior studies using the GDS-12D measure^13,28^. This aligns with a substantial body of evidence, and addresses a notable limitation^29^, in that results remained significant even when adjusting for comorbid symptoms and other confounds.

Additionally, this study provides novel understanding of the relationship between apathy and fatigue. In primary survival analyses, apathy and fatigue symptoms were separately associated with dementia and mortality. However, risk was attenuated when including both concurrently. We explored whether this could be due to confounding, by creating an apathy measure with one fatigue-related item removed. In sensitivity analyses using this measure, results were materially unchanged. This indicates that the fatigue-related GDS item was not the primary reason for shared variance between the GDS-3A and AQOL-12 measures. This contributes to an emerging picture of overlap between apathy and fatigue^21,22^. Further work is required to tease out their conceptual and methodological boundaries. Ours is the first study to suggest a longitudinal relationship between fatigue and subsequent dementia, to our knowledge, and aligns with those showing fatigue contributes to excess mortality^51^. Interestingly, fatigue appeared to be more relevant than apathy for health outcomes in the younger half of the cohort, and vice versa for the older group. However, caution should be taken in interpreting these findings, as they are based on a single-item fatigue measure, which did not capture its multidimensional conceptualisation, nor the worsening of symptoms after exertion (post-exertional malaise) which characterises primary fatigue syndromes^52^.

### Using different apathy measures modifies its association with dementia, but not mortality

In secondary analyses, we determined the impact of varying the apathy measure. Risk for dementia was apparent only when applying a strict cutoff to the self-reported apathy measure (GDS-3A), not a standard cutoff, nor when using a continuous score. This agrees with evidence of increased risk for those with ‘severe’, but not ‘moderate’, self-reported symptoms^44^. However, it contradicts another finding of a linear relationship^28^, and supports the need to determine the optimal cutoff for clinical significance on this GDS-3A measure, along with its other psychometric properties^30^.

Results were consistent between self- and informant-report measures in predicting mortality, but differed when incident dementia was the outcome. This aligns with very recent evidence that informant- but not self-reported apathy predicted β-amyloid and tau in unimpaired older adults^53^. However, informant-report measures of apathy have not been validated for use in pre-dementia populations^30^. Potentially, they capture only the most visible or severe cases of apathy, and might be more likely to result in significant findings, however these may not be representative for apathy as a whole. Results would also have been shaped by the psychometric characteristics of the measures used. The informant-report NPI apathy measure is commonly used, which allows for comparison between studies. However, its initial screening question leads to a floor effect in scores, and it lacks validity, according to recently reviewed high-quality evidence^30^. Together with the heterogeneity in meta-analyses caused by measurement issues^14,27^, these findings emphasize that different apathy measures, perspectives and scoring techniques lead to discrepant findings. Future research should focus on new measures designed for pre-dementia populations^54^, multiple perspectives and/or objective measures^55^.

### Future studies are needed to better understand the causal relationships that may underpin the observed associations between apathy and health outcomes

Mechanisms linking apathy with adverse health outcomes are not well-understood, and could be both causal and noncausal^24^. Our results support the hypothesis that apathy is a prodromal marker of dementia. This is in line with evidence linking apathy with neurodegeneration^53,56^, β-amyloid^57^ and tau^4^. Other proposed mechanisms include a common genetic predisposition^47^ and the vascular apathy hypothesis^58^, although in our sample, *APOE* L4 status and history of stroke or CVD did not interact with apathy, nor attenuate findings.

There might be also behavioural pathways between apathy and adverse outcomes. We have previously shown that apathy predicts lifestyle risks such as physical inactivity, as well as multimorbidity and poorer objective health^59^. Together, these factors may synergistically contribute to a greater risk for long-term outcomes such as mortality^60^. Early intervention could be critical in order to change this risk trajectory. Although no medications are currently approved for apathy, there are evidence-based interventions^56,61^, which could potentially help alleviate the daily struggle. It is unknown if intervening on apathy can help mitigate the future risk of dementia or mortality, but the current study warrants further attention to this topic.

### Strengths and limitations

Strengths of this research included the well-characterised population-based sample, and the availability of long-term outcome data. Dementia incidence over 12 years represents the longest follow-up in the apathy literature, and was determined by expert clinical consensus diagnosis, considered the gold standard. The use of two apathy measures, from different perspectives and with multiple cutoff scores applied, allowed novel comparisons and replication of primary results. Depression cases were included in primary analyses, and addressed using covariate adjustment, in comparison to most previous studies which excluded these cases.

Limitations included measurement issues for apathy and fatigue. Both can be considered multidimensional, long-term syndromes^5,52^, which may not have been fully captured by the brief measures available. Self-report symptoms were only required to be present during the week prior to baseline assessment. Assessing persistent symptoms (i.e. lasting ≥6 months, as per criteria for MBI^5^ and prolonged fatigue^20^) may allow for a more precise operationalisation^54^, although this was not possible using the MAS dataset. The fatigue item is from a quality of life questionnaire, and whilst it has convergent validity with longer fatigue questionnaires, it was not designed for diagnosis. As described earlier, the available apathy measures have evidence of suboptimal psychometric properties^30^. Both lack content validity, as they do not cover the emotional domain of the apathy syndrome. We attempted to address potential misclassification bias towards the null due to low sensitivity of the standard GDS-3A^37^ cutoff, by using a “strict” cutoff for primary analyses. Statistical power may have been reduced nonetheless. Reviews show the informant-report NPI apathy domain lacks construct validity^30^, hence we limited its use to secondary analyses. Additionally, generaliseability of the sample may be limited, due to the advantaged urban setting and lack of non-English speakers and cultural diversity. Selective dropout could have influenced our results for dementia, as participants without follow-up data had higher baseline apathy, depression and fatigue scores at baseline.

## Conclusion

This research illustrates the long-term health impacts of apathy and fatigue in the population. Individuals with apathy had elevated risk for dementia and mortality, independent of depression. However, the association with dementia was not clear-cut, suggesting reverse causality, whereas apathy was an independent risk factor for mortality.

## Data Availability

De-identified individual participant data collected during the Sydney Memory and Ageing Study are available to researchers who provide a methodologically sound proposal. Proposals should be directed to the Centre for Healthy Brain Ageing; to gain access, data requestors will need to sign a data access agreement.

## Consent Statement

All human participants provided informed consent.

## Author Contributions

Fleur Harrison: Funding acquisition, Conceptualisation, Methodology, Data curation, Formal analysis, Writing – Original Draft, Writing – Reviewing and Editing, Visualisation. Henry Brodaty: Supervision, Funding acquisition, Data curation, Writing – Reviewing and Editing. Moyra Mortby, Adam Guastella, Julian Trollor: Supervision, Writing – Reviewing and Editing. Perminder Sachdev: Funding acquisition, Data curation, Writing – Reviewing and Editing.

## Acknowledgements

The authors acknowledge the assistance of Dr Liesbeth Aerts in editing an early draft of the manuscript. The Sydney Memory and Ageing Study thanks the participants and their informants for their time and generosity in contributing to this research. We also acknowledge the dedication of the Sydney MAS research team: https://cheba.unsw.edu.au/research-projects/sydney-memoryand-ageing-study.

## Data sharing statement

De-identified individual participant data collected during the Sydney Memory and Ageing Study are available to researchers who provide a methodologically sound proposal. Proposals should be directed to the Centre for Healthy Brain Ageing; to gain access, data requestors will need to sign a data access agreement. The analytic code for this research publication will be available from the corresponding author, immediately following publication with no end date.

